# Socio-Cultural Factors and Experiences of School Going Teenage Mothers in Rural Zambia: A Phenomenological Study

**DOI:** 10.1101/2023.10.13.23296957

**Authors:** Dhally M. Menda, Rosemary K. Zimba, Catherine M. Mulikita, Mukumbuta Nawa, Jim Mwandia, Stephen F. Shamazubaula, Harrison Musonda, Karen Sichinga

## Abstract

**Background:** Teenage pregnancies disproportionately affect developing countries more than developed countries. This study aimed at exploring the social-cultural dynamics and experiences of teenage school-going mothers in a rural district of Eastern Province of Zambia to understand the lived experiences of teenage mothers in-depth.

**Methods:** This was a phenomenological study done in two schools in one of the provinces with the highest teenage pregnancy rates in Zambia. Data were analysed thematically in a deductive approach using a framework from a recent systematic review done in studies on teenage pregnancies from sub-Saharan Africa. A total of 26 school-going adolescent and young mothers were interviewed.

**Results:** The majority came from poor households, rural areas, lacked knowledge and access to sexuality education and contraceptive services before falling pregnant. Cultural beliefs and traditional practices fuel misconceptions on contraceptive and condom use, and create a narrative/expectation among the girls of wanting to experience sex. They experienced rejection and stigma during pregnancy and were now experiencing hardships in fending for themselves and their babies. Contemporary programs were either non-existent or not adequately addressing the economic and psychosocial challenges being faced by teenage mothers highlighting the need for more responsive interventions.

**Conclusion:** This study found that teenage and young mothers who re-entered school were more likely to be from poor and large families, first-borns and in day schools. Traditional beliefs and cultural practices influence They experience discrimination, rejection, low self-esteem and economic hardships.

**Strengths and Limitations of the Study:**

- The study was carried out in a rural district in one of the hard to reach areas in Zambia thus gives insights of the experiences of teenage and young mothers in a typical rural area where there is limited access to interventions such as comprehensive sexuality education and health services.
- To the best of the authors’ knowledge, this is the first time such a study has been done in Lundazi, a related previous study was done in Lusaka the capital of Zambia which is urban.
- A phenomenological study design gives the in-depth first hand experiences of what teenage and young mothers experience and go through before and during pregnancy, after delivery and when they re-enter school programs.
- The use of a framework that itemise personal, social, cultural, economic and health related characteristics of teenage and young mothers who re-enter school gives a holistic spectrum of factors that affect teenage and young women in school in rural areas.
- The main limitation is related to the design which is qualitative in nature and is therefore not representative of other rural areas or generalizable Zambia as a whole. There is therefore need for more research in other rural areas and other representative designs such as quantitative studies.

## Introduction

Teenage pregnancies and child marriages are among the drivers of maternal mortality globally (1, 2). Every year, there are about 21 million cases of teenage pregnancies reported globally, however, this problem disproportionately affects developing countries more than developed countries (1, 3, 4). The key drivers of teenage pregnancies in developing countries include poverty, lack of education, social and cultural practices, alcohol and substance abuse, limited access to health services and lack employment opportunities (4–7). Africa, particularly sub-Saharan Africa (SSA), has one of the highest rates of teenage pregnancies in the world (8–10). One study estimated the incidence of teenage pregnancy in sub-Saharan Africa at 143/1000 whilst in Europe it stood at 20/1000 girls aged 15 – 19 years (3). In Zambia, the latest national estimates of the prevalence of adolescent pregnancy were at 29% in 2018. However, there was a significant discrepancy between urban and rural areas, at 19 and 37%, respectively (11). Latest figures from the Ministry of General Education in Zambia (MoGE) indicate that a total of 15,222 school going girls got pregnant in 2017. One study done in Eastern Province estimated the prevalence of teenage pregnancies at 48% and child marriages at 13% (12). Eastern Province in Zambia has one of the highest rates of teenage pregnancies which was at 39.5% in 2018 compared to the national rate of 29.2% (11). The Zambian school system has 12 grades, with grades one to seven categorised as Primary School, and grades eight to twelve categorised as Secondary School; and the entry age in school is six to seven years. By the time a girl is in grade six or seven, she could attain menarche and if exposed to unprotected sex could become pregnant (13). The late adolescents’ age group of 15 to 19 years at which age the girls are in secondary school has been associated with teenage pregnancies (12, 14–16).

It is important to estimate the burden of teenage pregnancies in terms of prevalence and incidence, but this is only one aspect of the problem, as it does not put a ‘face’ to teenage mothers. Economic and socio-cultural factors influence the likelihood of a girl getting pregnant whilst still a teenager, and these may be missed in studies that focus on numbers such as prevalence and incidence (8, 10, 12, 17). Pregnancies occurring during the adolescent years have been associated with interference with the girl’s education. Studies have shown that about 30 – 60% of school dropouts among girls were due to teenage pregnancies or early marriages (4, 18, 19). The consequences of school dropout include poor employability, low earnings, poverty and poorer health outcomes among the girl child (3, 20, 21). Other than economic consequences, teenage pregnancies are also associated with increased morbidities such as haemorrhage, infections, miscarriages, hypertension, mental health problems, birth complications and death (12, 14, 22, 23). The Ministry of Education in Zambia introduced the school re-entry policy in 1997, for teenage mothers to continue their education after delivering (24, 25). While this policy has been implemented, its operationalization has often fallen short of its potential’ by being marred by challenges such as stigma, forced dropouts and failure to cope with added responsibilities (26). This present study aimed at exploring the social-cultural dynamics, economic and psychological experiences of teenage school-going mothers in a rural district of Eastern Province in Zambia, to further understand the lived experiences of adolescent mothers.

## Methods and Materials

### Study Design

This was a qualitative study that used a phenomenological approach. A phenomenological study helps to explore in-depth, the lived experiences of persons as told by themselves when they underwent a particular life experience, in this case, teenage pregnancy whilst at school (27–31). We chose this approach as it is more revealing in understanding some of the social circumstances that may have precipitated the girls to fall pregnant while at school, and their personal experiences during pregnancy and after delivery.

### Study Sites

The study was done in Lundazi District, in Eastern Province of Zambia, it is situated about 750 kilometres from Lusaka the capital of Zambia. It was purposively selected because Lundazi is a rural district located in a province with one of the highest teenage pregnancies in the country. Two schools were selected, namely Lusunta Secondary and Lundazi Day secondary schools, because they were the only ones who implemented the school Re-entry Policy in the district.

### Study Population and Sampling

A list of all school-going teenage and young mothers aged 15 – 24 years at Lusunta and Lundazi Day Secondary Schools was obtained from the school authorities. The respondents were purposively sampled to obtain maximum variation in terms of age, grade and social backgrounds. A total of 26 interviews were done, and thematic saturation was reached.

### Data Collection

In-depth interviews were done with individual respondents, using guides that had open-ended questions and a recording of the interviews was done. Two researchers did the interviews while two assistants recorded and took notes in two separate groups. Each interview took thirty minutes to one hour and allowed for further clarifications outside the initial interviews. The data was transcribed from the interview notes and recordings into Microsoft word by the two assistants who took part in the interviews. To ensure credibility and trustworthiness of the data, the two researchers quality checked all the manuscripts, triangulating with the recordings and field notes before analysis.

### Data Analysis

The data was analysed using thematic analysis in N-Vivo software version 9 (32) in a deductive approach using a model proposed by a systematic review on adolescent pregnancies in sub-Saharan Africa (9) which suggested individual, socio-cultural, environmental, economic and health-related factors as determinants of teenage pregnancies. We identified the issues that were mentioned and grouped them under a common theme by two researchers working independently and consensus was reached. Where an agreement was not reached, a third person was used as a tie-breaker. The themes were further analysed into subthemes and reported narratively in a logical manner. Translated verbatim quotations were written alongside the synthesised data. Basic socio-demographic variables were analysed quantitatively in SPSS Software version 22 (33), and summary statistics were presented to give a contextual perspective of respondents to the readers.

### Ethical Considerations

The rights, dignity, welfare, respect and courtesy of the participants were respected in this study. The provincial, district and school authorities were informed about the study, and they gave permission. The parents/guardians gave informed consent for all girls who were below 18 years, and the girls themselves also gave assent. The girls who were 18 years and above gave informed consent before the interviews were done. Participants were allowed to decline or change their mind and discontinue the interview if they so wished. The names of participants were kept anonymous and pseudo names were used. ERES Converge Ethics Committee cleared the protocol for the study (Protocol No. 2020-Nov-003).

## Results

### Basic Description of Respondents

A total of 26 school going adolescents and young mothers who had re-entered school after pregnancies were interviewed in the study. The majority of the respondents resided in rural areas 20 (77%) while 6 (23%) lived in urban areas. All respondents were in senior secondary school at the time of the interviews. The median age was 19 years, with an interquartile range (IQR 18 – 20), the youngest among the respondents was 17 years old, whilst the oldest was 22 years old. Of these 13 (50%) were in Grade ten, 10 (39%) in grade eleven and 3 (11%) in grade twelve.

In terms of socio-economic status, proxy indicators were used such as ownership of high-value assets, within the Zambian context like bicycles, refrigerators and television sets. Of these, only 1 (4%) came from a family that had a bicycle, refrigerator and a television set, 7 (27%) had either a bicycle or a television set, and the majority 18 (69%) came from families that did not own any of the assets. In terms of highest educational attainments of the male guardians for the girls, the majority 19 (73%) of the girls had male guardians who had only reached either primary (grades 1 – 7) or junior secondary/high school (grades 8 and 9). Only one girl had a male guardian who had reached tertiary education. In term of female guardians, 15 (58%) had female guardians who had either primary or junior secondary/high school, 1 (4%) had a guardian who had reached senior secondary school, and 8 (31%) had a female guardian who had not attended formal schooling at all. Also, the majority 21 (81%) of the girls came from families with more than four children, only 5 (19%) came from families with three or fewer children. Also, it was found out that the highest proportion of the girls 10 (38%) were first-borns in their families. Another interesting finding was that 23 (88%) of the girls had undergone initiation ceremonies, whilst three did not undergo the processes of initiation ceremonies.

### Individual Factors

Individual factors contributed to the girls getting pregnant such as their age. The girls were in the adolescent age group; they had attained menarche and were physiologically ready for sexual activities and pregnancy. Many reported having an attraction for the opposite sex and having sexual desires as exemplified in this excerpt:

*‘I just wanted to experience it; I had feelings and desire’* (Female Respondent, age range 15 - 18 years).

The median age of menarche among the respondents was 13 years (IQR 12 – 14 years) while the median age of sexual debut was 16 years with (IQR 15 – 17 years). By the age of 18 years, 92% of our respondents already had had their first sexual experience. The median age at first pregnancy among our respondents was 17 years (IQR 16 – 18 years), by the age of 18 years, about 88% of them already had had their first pregnancy. This indicates that within a year of starting sexual activities, the majority of our respondents had fallen pregnant. The median school grade attended at the time of first pregnancy was grade 10 (IQR 9 – 11), by the time they finished grade 11, 92% of the girls had had their first pregnancy. The participants also reported practising safe/protected sex 17 (65%). However, this was after they had given birth. They had learnt about safe sex later on, after their pregnancies. Most of the teen mothers were now knowledgeable about the use of condoms to protect against unwanted pregnancies and also prevent the spread of sexually transmitted infections (STIs). The following are some of the verbatim quotations from the respondents, concerning safe sex:

*‘Safe sex is having protected sex using a condom.’* (Female Respondent, Age range 19 - 21 years)

*‘To protect oneself from pregnancy and diseases such as HIV/AIDS, STIs and Syphilis’* (Female Respondent, Age range 15 - 18 years old).

The type of school that the girls attended also mattered; 58% of them were day scholars when they first got pregnant, 35% were weekly boarders, and only 7% of them were in a boarding school. In other words, the odds ratio of falling pregnant among the respondents, when one was in a boarding school was 0.05 (95% CI 0.02 – 0.13) compared to being in a day school, implying that boarding school cut the odds by about 95%. This study did not elicit data on the use of alcohol and substance abuse which has also been associated with increased vulnerability to sexual intercourse and pregnancies.

### Sociocultural, Environmental and Economic Factors

#### Peer Pressure and Poverty

Some of the social factors reported by the respondents included peer pressure among the girls to have sex. Such kind of pressure would make the girl conform to what is socially acceptable with her friends. Other girls reported the lack of basic needs as a driver for them to engage in sexual activities. Below is are excerpts from some of the respondents:

*‘You won’t have anything in your life when you don’t have a boyfriend and also when you refuse the demands of your boyfriend’* (Female Respondent, Age range 15 - 18 years old).

‘*I lacked support in basic needs from the family such as food, clothes, lotion and soup*’ (Female Respondent, Age range 15 - 17 years old).

#### Lack of Parental guidance

Many of the respondents attributed their being sexually active to lack of information or knowledge on sexual matters.

*‘Lack of communication with the parents on sexual issues as they are said to be taboo. My parents never talked to me about issues of sexual intercourse’* (Female Respondent, Age range 19 - 21 years old)

#### Lack of Knowledge

Many respondents attributed their being sexual active and not using condoms or contraceptives to lack of information or knowledge. They learnt more about safe sex after they got pregnant during antenatal visits when they had more contact with health care workers.

*‘Lack of information because I didn’t know what I was doing until I got pregnant’ (*Female Respondent. Age range 19 - 21 years old).

‘*I learnt more about contraceptive methods during antenatal visits when I was pregnant, before that, I did not know a lot*’ (Female Respondent, Age range 22 - 25 years old).

### Traditional Practices and Cultural Beliefs

Respondents reported on some traditional practices such as initiation ceremonies for girls which gives them a rite of passage into adulthood. The initiators teach the girls how to have sex with a man. Others reported on cultural beliefs that a girl would be viewed as ‘normal’ if only she had a boyfriend as exemplified below:

*“At a certain age you need to have a boyfriend….sex is nice and needs to be practised”* (Female Respondent, Age range 15 - 18 years old)

Other factors that were raised by the respondents which included the following: Family breakdown, desire to have a sexual experience, school far from home, the glamorisation of sexual intercourse and boy/girlfriend issues by others, coercion by a boyfriend to have sex otherwise you do not love him and acceptance of early sex by the community:

*‘If I don’t have sex with him, it means I don’t love him, and he won’t give me the money’* (Female Respondent, Age range 15 - 18 years old)

### Health Service Related Factors

Some factors, such as the provision of knowledge and access to condoms and contraceptive methods, were reported as having been lacking among the girls before they got pregnant. Some only got to know about these during antenatal visits and after delivery. Lack of sexuality education and misconceptions on contraceptives were also reported. Others thought that contraceptives were only for married people and that it could cause permanent infertility if one has not given birth before, as quoted below:

‘*they (contraceptives) destroy you and you cannot have a child again in the future*’ (Female Respondent, Age range 15 - 18 years old).

Other misconceptions were that contraceptives could increase the menstrual length and cause heavy bleeding, and this rumour made the schoolgirls apprehensive about them. Once the girls had adequate contact with health care workers at antenatal and under-five clinics, they reported positive attitudes towards contraceptives, indicating a need for sexual and reproductive health education in schools to girls, before they fall pregnant. Similarly, there were misconceptions that only males had a say on whether to use a condom or not during sexual intercourse. Others felt that it was awkward for a woman to carry condoms, as it may show that she is promiscuous:

‘*Only men are allowed to carry condoms, and it is not normal for ladies to be found with condoms. I was also shy to collect condoms from the clinic*’ (Female Respondent, Age range 15 - 18 years old).

### Experiences during Pregnancy

In addition to the exploring for some factors that drive teenage pregnancies among our respondents, the study also explored the lived experiences of adolescent mothers during their pregnancies. Their experiences included, among many others, hostile and aggressive reactions of parents/guardians, male folks refusing their responsibility in recognising the pregnancy, sickness and body pains, lack of concentration at school, and lack of basic needs to cater for the mother and the baby’s needs:

*‘My mother disowned me, and she stopped talking to me. My boyfriend refused responsibility…lack of concentration at school, panic and psychological stress’* (Female Respondent, Age range 15 - 17 years old).

Some other experiences reported included, being chased from home, contemplating abortion, suicidal thoughts, lack of family support, abandonment by close friends, mockery, discrimination, lack of preparation for motherhood, and also lack of support from the boys/men responsible of the pregnancy.

*‘I was psychologically disturbed and became shy of moving, sickness of the eyes, swelling of legs and shortage of blood, the preparations for the baby were difficult.’* (Female Respondent, Age range 19 - 21 years old)

### Support Needed by Teenage Mothers

Almost all the respondents pleaded for support from their families, friends, school, church and the government, stating that they were still human beings. The following were among the typical kinds of support these teen mothers wanted during pregnancy, during their babies’ care, as well as when back to school: psychological support, encouragement to continue with school even after falling pregnant, sponsorship, helping them with the care of the baby, material support for their babies, e.g. hampers, clothes, food, toiletries, etc., sexual and reproductive health education and services, needs for inclusion and closeness with other family members and friends, and giving them a second chance at school.

*‘No support from the guy, I stopped school leading into a delay…So I started doing piece work, and it was painful that I was pregnant’* (Female Respondent, Age range 19 - 21 years old).

### Lessons Learnt from Experiences of Pregnancy

Most of the participants reported that they would not want to fall pregnant again while at school, making references to some of the unpleasant experiences that they went through during the pregnancies and afterwards. Most of them were more ambitious for excellence and development than before.

*‘I wouldn’t be pregnant again because of consequences of pregnancy that I experienced such as missing school where I was not going to school, problems of upkeep of the child and also lack of basic needs’* (Female Respondent, Age range 22 - 25 years old).

*‘Teenage pregnancy is bad for teenagers because bones are not strong leading to operations, hence, it is very dangerous we just need to abstain’* (Female Respondent, Age range 19 – 21 years old).

Others cited having gained more knowledge of how pregnancies come about as well as increased knowledge on the use and access to both contraceptives and condoms (family planning). Abstinence was another tool that was mentioned by many teen mothers to be the way forward in order not to become pregnant again in the future without proper planning.

‘*I learnt more about contraceptive methods during antenatal visits when I was pregnant such as condoms, oral and injectable contraceptives*” (Female Respondent, Age range 22 - 25 years old).

## Discussion

This study, as part of the baseline assessment of the AGAPE project under CHAZ, was able to profile adolescent girls and young women who had fallen pregnant while at school and gave their experiences before, during pregnancy and after delivering. The girls were mainly from rural areas, from poor families that did not own assets, had guardians who are not educated, big families and had undergone traditional initiation ceremonies. The study showed that school-going girls are in the adolescent age group and experience personal, socio-cultural, environmental, and economic and health services factors that predispose them to teenage pregnancies as reported by other studies (9, 12, 34). The average age of first pregnancy in our study was 17 years, in contrast to developed countries such as the United Kingdom where it was 27 years (35).

There are some personal changes such as puberty and menarche, driven by hormonal changes, which also trigger sexual feelings among the girls (36, 37). Program interventions in sexuality education recognise this and need to empower the girls to make informed decisions on their sexuality to abstain or protect themselves if they are going to engage in sexual activities (34, 38, 39). Other than personal biological needs that have to be considered, in rural set-ups such as Lundazi district in Zambia, there are other socio-cultural factors that can still drive adolescent girls to engage in unsafe sexual practices such as initiation ceremonies as highlighted in our study. Poverty, peer pressure, and cultural beliefs and personal desires among others as has been elicited in this study can lead to teenage pregnancies, complications of pregnancy and sexually transmitted infections. These findings have also been corroborated by other researchers (40, 41), including in a systematic review of studies in Sub-Saharan Africa (9). Some researchers however, counteract this and argue that empowered women and those with higher education were significantly associated with risky sexual behaviour (42).

One interesting finding in this study was that firstborn girls were more likely to contract teenage pregnancies compared to non-first born. This finding was also found in a study in western Nigeria (43). The reasons may not be clear but probably because the firstborn girls, particularly from poor households, may try to fend for themselves and other siblings (44). In addition, environmental factors such as attending day schools and walking long distances to school were shown to be associated with a higher risk of contracting teenage pregnancies compared to boarding schools. This finding has also been supported in other studies carried out in sub-Saharan African countries like South Africa and Ghana (14, 45, 46). Economic factors have also been shown to affect teenage pregnancies. Girls from poor households fall prey to men who are able to give them gifts to meet their personal needs and some fancied things like clothes and cell phones to suit with their peers (47–49). While our data showed poverty as one of the drivers, evidence from developed countries such as New Zealand and the UK indicated personal instability, hardy upbringing and minority neighbourhoods as some of the key drivers of teenage pregnancy (50, 51).

Health services in rural areas in developing countries fall below other areas such as urban areas; conversely, access to reproductive health services such as appropriate information and commodities may be compromised. The most rural districts such as Lundazi have actually been noted to have higher teenage pregnancies relative to more urban settings (52). Rural areas also experience a double-edged sword of limited services as well as abounding traditional beliefs and practices. Some traditional practices in rural such as initiation ceremonies train teenage girls in how to handle men in bed which elicit wanting to experience what one has been taught (53). Some communities actually accept and consider early child-bearing as a norm (37).

Teenage mothers also reported feelings of rejections from family and friends, including stigmatisation by society, these findings were also reported by other researchers in both developed and developing countries (54–56). They further desired support instead of ridicule and stigmatisation. These findings are important to foster safer environments for the teenage mothers to avoid depression, infanticide or suicide (57, 58). Further, the negative feelings of teenage pregnancy were also associated with a feeling of wanting to abort the pregnancy; this, too, was corroborated by other researchers (59). Supportive environments are, therefore, more desirable to teenage pregnancies and teenage mothers for the safety of the teenager and the baby (60, 61). Additionally, adolescent mothers also reported requiring material incentives for them to remain in school because of the increased responsibilities, the requirement for support was also reported by other studies from both developing and developed countries (21, 62).In developed countries, vulnerable teenage mothers receive support from government welfare departments such as a living stipend, food stamps which they can redeem at local supermarkets, health insurance and social support such as counselling (21, 63). Some African countries such as South Africa also give a social grants to teenage mothers which has shown to help teenage mothers regain traction (61, 64). Some teenage mother seems to have emerged more ambitions and determined to succeed in education despite going through the pregnancy episode. This finding was also corroborated by studies done in schools in the Africa region where adolescent mothers who re-enter schools develop resilience and will to compete school despite the pregnancy experience (61, 65).

## Conclusion

This study which aimed at exploring the lived experiences of teenage mothers in a district in one of the provinces with the highest Teenage pregnancies in Zambia. Teenage pregnancy was found to be associated with rural poor girls who had low access to comprehensive sexuality education and contraceptive services. Further, socio-demographic and cultural practices such as initiation ceremonies tended to create expectations and desires to engage in sexual activities among the adolescent girls. In addition, environmental factors in rural areas such as walking long distances to school were also identified as influencing factors. Once pregnant, adolescent girls experienced reprisals during pregnancy and economic hardships upon re-entering schools and need material and psychological support to continue with their school due to increased demands. While governments in developing countries and some emerging economies in Africa are able to supplement vulnerable teenage mothers with a living stipend and other social services such as counselling and medical insurance, this is not the case with Zambia and it was highlighted as one of the requirements by the teenage mothers. Programs such as AGAPE that provide comprehensive sexuality education, pay school fees for the girls and offer a living stipend for vulnerable girls are addressing some of the highlighted needs. The program may reduce the incidences of teenage pregnancies as well as providing safer learning environments for adolescent and young mothers who re-enter school.

## Data Availability

Data is available upon request to the corresponding author

## Acknowledgements

The Authors would like to sincerely thank CHAZ management, the donor particularly the Global Fund to fight AIDS, Tuberculosis, and Malaria (GFATM), the Ministry of Education (MoE) at headquarters and provincial level in Eastern Province, the teachers at the selected two schools and the respondents.

## Authors Contributions

All authors contributed in the conception of the research and development of the protocol, DMM, RKZ, SFM, CMM, JM & HM collected the data, DMM & NM drafted the manuscript, DMM & KS provided the overall leadership. All authors provided the reviewed the manuscript and approved it before submission for publication. DMM Dhally Mutombo Menda, NM Nawa Mukumbuta, RKM Rosemary Kalumiana Zimba, SFM Stephen F. Shimaubaula, CMM Catherine Mukuka Mulikita, JM Jim Mwandia, HM Harrison Musonda, KS Karen Sichinga.

## Data Availability Statement

The original datasets are available at the CHAZ Secretariat upon request through email by contacting the Executive Director on.

